# Identification of potential changes in protein abundance associated with post-traumatic stress disorder through brain proteome-wide association study

**DOI:** 10.1101/2025.08.28.25334625

**Authors:** Jiewei Liu, Zhongchun Liu

**Affiliations:** Department of Psychiatry, Wuhan Mental Health Center, Wuhan 430012, Hubei, China; Wuhan Hospital for Psychotherapy, Wuhan 430012, Hubei, China; Department of Psychiatry, Renmin Hospital of Wuhan University, 238 Jiefang Road, Wuhan, 430060, Hubei, China; Taikang Center for Life and Medical Sciences, Wuhan University, 115 Donghu Road, Wuhan, 430071, Hubei, China

**Author notes:** Correspondence to: Jiewei Liu, Wuhan Mental Health Center, Wuhan, 430022, Hubei, China.; Zhongchun Liu, Renmin Hospital of Wuhan University, Wuhan 430060, Hubei, China.

## Abstract

**Aim:** Post-traumatic stress disorder (PTSD) is a common mental disorder with a substantial genetic background. Recent genome-wide association analysis (GWAS) has identified 95 genome-wide significant (GWS) loci in a large cross-ancestry cohort comprising over 1 million samples. However, bridging the GWS findings to potential therapeutic candidates is lagged.

**Method:** In this study, we integrated the most up-to-date PTSD GWAS data with two human brain proteome datasets using FUSION software to identify potential changes in protein abundance that confer risk for PTSD through proteome-wide association analysis (PWAS).

**Results:** We identified 20 and 24 proteome-wide significant (PWS) genes from the Banner (n=152) and ROSMAP (n=376) PWAS results, yielding a total of 33 non-overlapping PWS genes. Notably, *CTNND1* was the top PWS gene in both PWAS analysis panels. In addition to *CTNND1*, the genes *TMEM106B*, *ICA1L*, *KHK*, *GPX1*, *CCBL2*, *MKRN1*, *INPP4A*, *SIRPA*, *CACNA2D2*, and *PDLIM2* also reached PWS levels in both Banner and ROSMAP proteome datasets. Furthermore, we performed colocalization, drug-gene interaction analysis, and synapse function annotation analysis.

**Conclusion:** Our study uncovered the changes in protein abundance that underlie PTSD etiology, and these genes represent promising candidates for further exploration of the mechanisms underlying PTSD pathogenesis.

## Introduction

Posttraumatic stress disorder (PTSD) is a common psychiatric disorder that develops in individuals who have experienced traumatic events. The clinical symptoms of PTSD include intrusive thoughts, avoidance behaviors, negative cognition and mood symptoms, as well as changes in arousal and reactivity [1]. It has been reported that the prevalence of PTSD is approximately 3.9% in the general population, increasing to 5.6% among those who have experienced trauma [2]. The high prevalence of PTSD results in a significant economic burden on society [3]. Therefore, effective and efficient methods are needed for the treatment of PTSD. While environmental factors play an important role in PTSD, genetic background should not be overlooked. Recent studies estimate that the heritability of PTSD is about 46% [4]. Genome-wide association studies (GWASs) are commonly used strategies for dissecting the genetic basis of complex diseases/traits, including many neuropsychiatric disorders. Several GWASs have been conducted to explore genomic risk loci associated with PTSD [5–7]. Recently, the Psychiatric Genomic Consortium for PTSD (PGC-PTSD) performed the largest PTSD GWAS to date, which included 1,307,247 samples from 88 studies and reported 95 genome-wide significant (GWS) loci. Most GWS variants are located in non-coding regions, it is implied that these variants may contribute to disease risk by regulating gene expression rather than directly affecting protein function [8]. However, bridging the summary statistics from PTSD GWAS with disease risk genes is a key issue for interpreting the loci identified in GWAS, due to the complex linkage disequilibrium structure of the human genome [9].

Multi-omics integration methods, such as transcriptome-wide association analysis (TWAS) and Summary-data-based Mendelian randomization (SMR) [10, 11], have been developed to identify candidate risk genes for diseases/traits by integrating GWAS summary statistics with transcriptomic or proteomic data. In this study, we conducted a brain proteome-wide association study (PWAS) for PTSD by combining the new PGC released PTSD GWAS summary statistics and two brain proteome datasets. Our goal was to identify changes in protein abundance related to PTSD, which will be helpful for further studies on disease mechanisms and therapeutic target evaluation.

## Methods

### PTSD GWAS summary statistics

Nievergel et al. (2024) conducted a multi-ancestry PTSD GWAS with the largest sample size to date (n=1,307,247) [12], including three ancestries: European, African, and Latin American. In this study, we downloaded the European ancestry GWAS summary for the subsequent PWAS analysis (n=1,222,882). For detailed information about the PTSD GWAS data, please refer to the original GWAS publication.

### Brain proteome datasets

We retrieved two brain proteome datasets, both containing protein abundance data and genotype information from a previous study (https://www.synapse.org/Synapse:syn23627957) [13]. The first dataset is from the Religious Orders Study and Rush Memory and Aging Project (ROSMAP) (n=376), and the second dataset is from Banner Sun Health Research Institute (Banner, n=152). Both ROSMAP and Banner brain proteomes were generated from postmortem samples of the brain dorsolateral prefrontal cortex (dlPFC) region, with protein abundance measured using isobaric tandem mass tag (TMT) peptide labeling. Genotypes were generated using various methods, including whole genome sequencing (WGS) and whole genome genotyping arrays (e.g., Illumina and Affymetrix platforms). All samples are of European descent.

### PWAS analysis by FUSION software

We performed PWAS analysis using FUSION software (https://gusevlab.org/projects/fusion/) [10]. The brain proteome weigh files for both Banner and ROSMAP were downloaded from https://www.synapse.org/#!Synapse:syn23627957 [13]. We used the default parameters and 1000 Genome project European samples as the reference for FUSION analysis. Additionally, we employed the –coloc_P tag to trigger colocalization analysis.

### Drug-gene interaction analysis

Drug-gene interactions were analyzed using DGIdb databases (https://www.dgidb.org/) [14]. The DGIdb database curates the drug-gene interactions derived from well-established databases such as DrugBank and ChEMBL, as well as through text mining from literature. DGIdb is an ongoing project, with the latest version was DGIdb 5.0. It contains approximately 70,000 drug-gene interactions, including over 10,000 genes and 20,000 drugs.

### Gene set enrichment analysis in synapse

To explore the potential functions of the PWS genes, we utilized SynGO online web tool (https://www.syngoportal.org/) for annotation [15]. The newly released SynGO 1.2 includes 4,218 annotations of a total of 1,620 genes. The annotations and genes are curated by experts from the SynGO consortium. For a more detailed description, please refer to the SynGO paper

## Results

### 33 genes reached proteome-wide significant (PWS) level in PWAS analysis

We integrated the PTSD GWAS with two brain proteomes to conduct a PWAS. In the ROSMAP PWAS analysis, we identified 24 PWS genes, of which 15 replicated their association with PTSD in the Banner PWAS analysis (**Figure 1a**, **Table 1**). Additionally, 20 PWS genes were identified in the Banner PWAS analysis, with 14 replicating in the ROSMAP PWAS analysis (**Figure 1b**, **Table 1**). 11 genes, including *CTNND1*, *TMEM106B*, *ICA1L*, *KHK*, *GPX1*, *CCBL2*, *MKRN1*, *INPP4A*, *SIRPA*, *CACNA2D2*, and *PDLIM2*, exhibited PWS in both PWAS analysis panels (**Figure 1**). Notably, *CTNND1* was the most significant gene in both the ROSMAP (*P value* = 1.68×10^-17^) and Banner *(P value* = 8.05×10^-13^) PWAS results. In total, we identified 33 non-overlapping PWS genes in our PWAS analysis.

**Figure 1.**
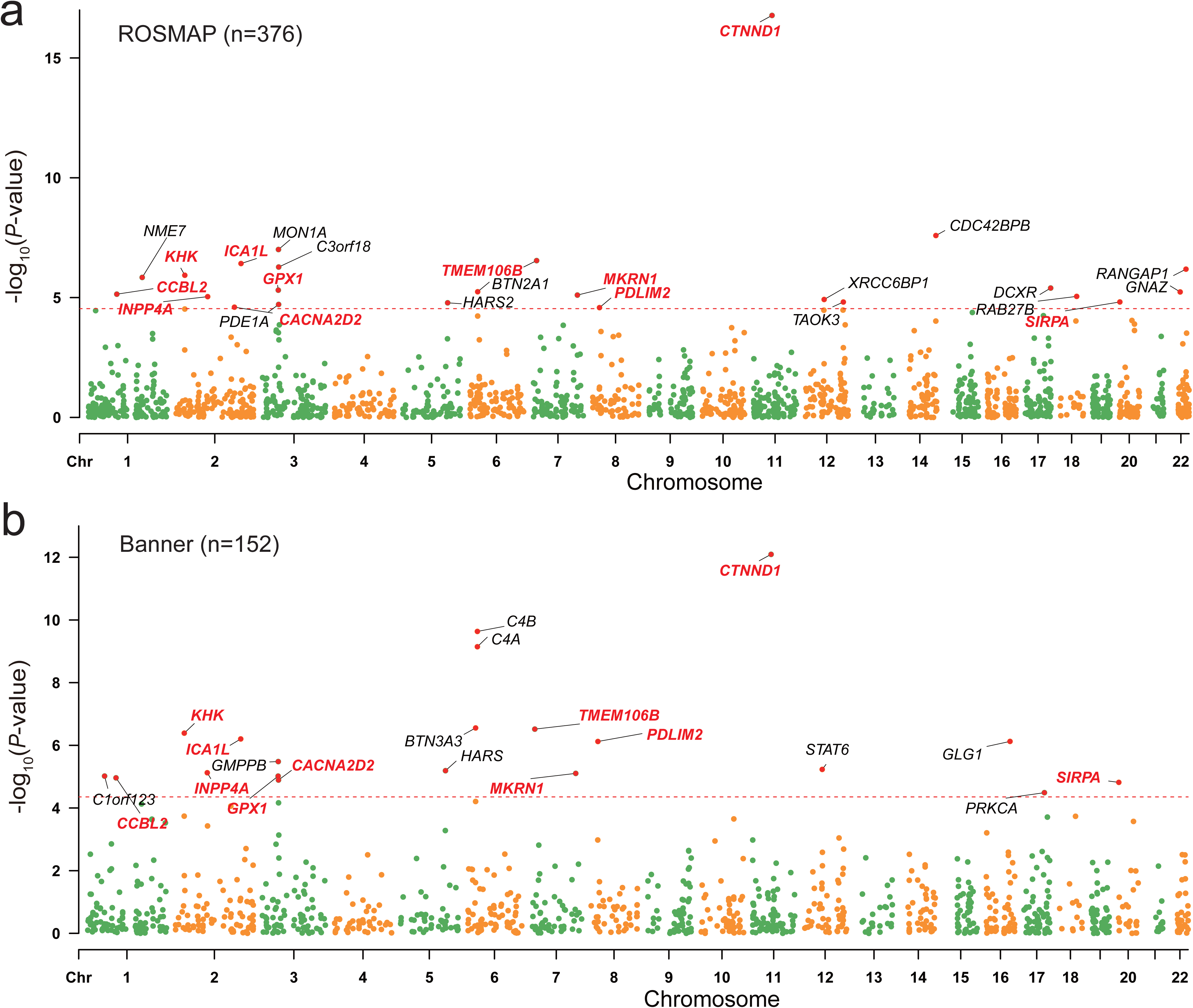
The Manhattan plot of the brain proteome-wide association study (PWAS) analysis. a) Results of the PWAS analysis integrating ROSMAP (n=376) proteome data with PTSD GWAS summary statistics. The red dashed line indicates the Bonferroni-corrected significance level (*P* = 2.89×10^-05^); b) Results of the PWAS analysis integrating Banner (n=152) proteome data with PTSD GWAS summary statistics. The red dashed line indicates the Bonferroni-corrected significance level (*P* = 4.43×10^-05^).

**Table 1.** Proteome-wide association studies result of PTSD GWAS and two brain proteome data.

| Gene* | Chromosome | ROSMAP |  | Banner |  | replicated |
| --- | --- | --- | --- | --- | --- | --- |
|  |  | P value | COLOC PP4 | P value | COLOC PP4 |  |
| **<u>CTNND1</u>** | 11 | $1.68 \times 10^{-17}$ | 0.931 | $8.05 \times 10^{-13}$ | 0.533 | Yes (Both meet PWS) |
| *CDC42BPB* | 14 | $2.57 \times 10^{-08}$ | 0.54 | NA | NA | NO |
| *MON1A* | 3 | $9.85 \times 10^{-08}$ | 0.494 | NA | NA | NO |
| **<u>TMEM106B</u>** | 7 | $2.86 \times 10^{-07}$ | 0.989 | $3.04 \times 10^{-07}$ | 0.989 | Yes (Both meet PWS) |
| **<u>ICA1L</u>** | 2 | $3.79 \times 10^{-07}$ | 0.987 | $6.25 \times 10^{-07}$ | 0.986 | Yes (Both meet PWS) |
| *C3orf18* | 3 | $5.31 \times 10^{-07}$ | 0.89 | $6.84 \times 10^{-05}$ | 0.757 | Yes |
| *RANGAP1* | 22 | $6.50 \times 10^{-07}$ | 0.509 | NA | NA | NO |
| **<u>KHK</u>** | 2 | $1.17 \times 10^{-06}$ | 0.932 | $4.08 \times 10^{-07}$ | 0.953 | Yes (Both meet PWS) |
| *NME7* | 1 | $1.44 \times 10^{-06}$ | 0.722 | $7.43 \times 10^{-05}$ | 0.52 | Yes |
| *DCXR* | 17 | $4.00 \times 10^{-06}$ | 0.013 | NA | NA | NO |
| **<u>GPX1</u>** | 3 | $4.90 \times 10^{-06}$ | 0.916 | $9.60 \times 10^{-06}$ | 0.908 | Yes (Both meet PWS) |
| *BTN2A1* | 6 | $5.70 \times 10^{-06}$ | 0.043 | $6.19 \times 10^{-05}$ | 0.057 | Yes |
| *GANZ* | 22 | $5.81 \times 10^{-06}$ | 0.312 | NA | NA | NO |
| **<u>CCBL2</u>** | 1 | $7.19 \times 10^{-06}$ | 0.801 | $1.09 \times 10^{-05}$ | 0.839 | Yes (Both meet PWS) |
| **<u>MKRN1</u>** | 7 | $7.86 \times 10^{-06}$ | 0.929 | $7.86 \times 10^{-06}$ | 0.787 | Yes (Both meet PWS) |
| *RAB27B* | 18 | $8.91 \times 10^{-06}$ | 0.908 | $1.84 \times 10^{-04}$ | 0.786 | Yes |
| **<u>INPP4A</u>** | 2 | $9.13 \times 10^{-06}$ | 0.804 | $7.49 \times 10^{-06}$ | 0.205 | Yes (Both meet PWS) |
| *XRCC6BP1* | 12 | $1.20 \times 10^{-05}$ | 0.762 | NA | NA | NO |
| **<u>SIRPA</u>** | 20 | $1.52 \times 10^{-05}$ | 0.875 | $1.52 \times 10^{-05}$ | 0.875 | Yes (Both meet PWS) |
| *TAOK3* | 12 | $1.53 \times 10^{-05}$ | 0.976 | 0.575 | NA | NO |
| *HARS2* | 5 | $1.65 \times 10^{-05}$ | 0.504 | NA | NA | NO |
| **<u>CACNA2D2</u>** | 3 | $1.93 \times 10^{-05}$ | 0.436 | $1.28 \times 10^{-05}$ | 0.292 | Yes (Both meet PWS) |
| *PDE1A* | 2 | $2.51 \times 10^{-05}$ | 0.517 | NA | NA | NO |
| **<u>PDLIM2</u>** | 8 | $2.62 \times 10^{-05}$ | 0.867 | $7.54 \times 10^{-07}$ | 0.451 | Yes (Both meet PWS) |
| *C4B* | 6 | NA | NA | $2.32 \times 10^{-10}$ | 0.2 | NO |
| *C4A* | 6 | NA | NA | $7.14 \times 10^{-10}$ | 0.882 | NO |
| *BTN3A3* | 6 | $5.89 \times 10^{-05}$ | 0.04 | $2.79 \times 10^{-07}$ | 0.936 | Yes |
| *GLG1* | 16 | NA | NA | $7.50 \times 10^{-07}$ | 0.031 | NO |
| *GMPPB* | 3 | $2.91 \times 10^{-04}$ | 0 | $3.30 \times 10^{-06}$ | 0 | Yes |
| *STAT6* | 12 | NA | NA | $5.87 \times 10^{-06}$ | 0.772 | NO |
| *HARS* | 5 | $8.63 \times 10^{-02}$ | NA | $6.44 \times 10^{-06}$ | 0.347 | NO |
| *C1orf123* | 1 | $1.18 \times 10^{-03}$ | 0.079 | $9.57 \times 10^{-06}$ | 0.674 | Yes |
| *PRKCA* | 17 | $8.04 \times 10^{-02}$ | NA | $3.24 \times 10^{-05}$ | 0.001 | NO |

\* Genes that with proteome-wide significant (PWS) level are shown. Genes that shown PWS in both proteome datasets are in bold and undelined.

### Colocalization analysis further confirmed the potential causal role of 23 PWS genes

To confirm the potential causal relationship between the PWS genes and PTSD, we conducted a colocalization analysis. Among the 33 PWS genes, 23 exhibited a PP4 value greater than 0.5 in at least one PWAS analysis panel (**Table 1**). *TMEM106B*, *ICA1L*, *KHK*, *GPX1*, *CCBL2* and *SIRPA* demonstrated high PP4 values (>0.8) in both PWAS panels. *CTNND1* showed a high PP4 in the ROSMAP PWAS analysis (*PP4*=0.931) but a relatively low value in the Banner PWAS result (*PP4*=0.533).

### Drug target gene analysis identified multiple drug-gene interactions of PWS genes

To investigate potential therapeutic drugs targeting the PWS proteins identified in this study, we utilized DGIdb to explore the drug-gene interactions. We found 12 genes with documented interactions (**Table 2**), including *PDE1A* (2 interactions)*, C4A* (4 interactions)*, C4B* (4 interactions)*, PRKCA* (24 interactions)*, GPX1* (2 interactions)*, TAOK3* (1 interactions)*, CACNA2D2* (14 interactions)*, STAT6* (15 interactions)*, KHK* (1 interactions)*, SIRPA* (1 interactions)*, CTNND1*(11 interactions)*, and GLG1* (5 interactions) in the DGIdb databases.

**Table 2.**
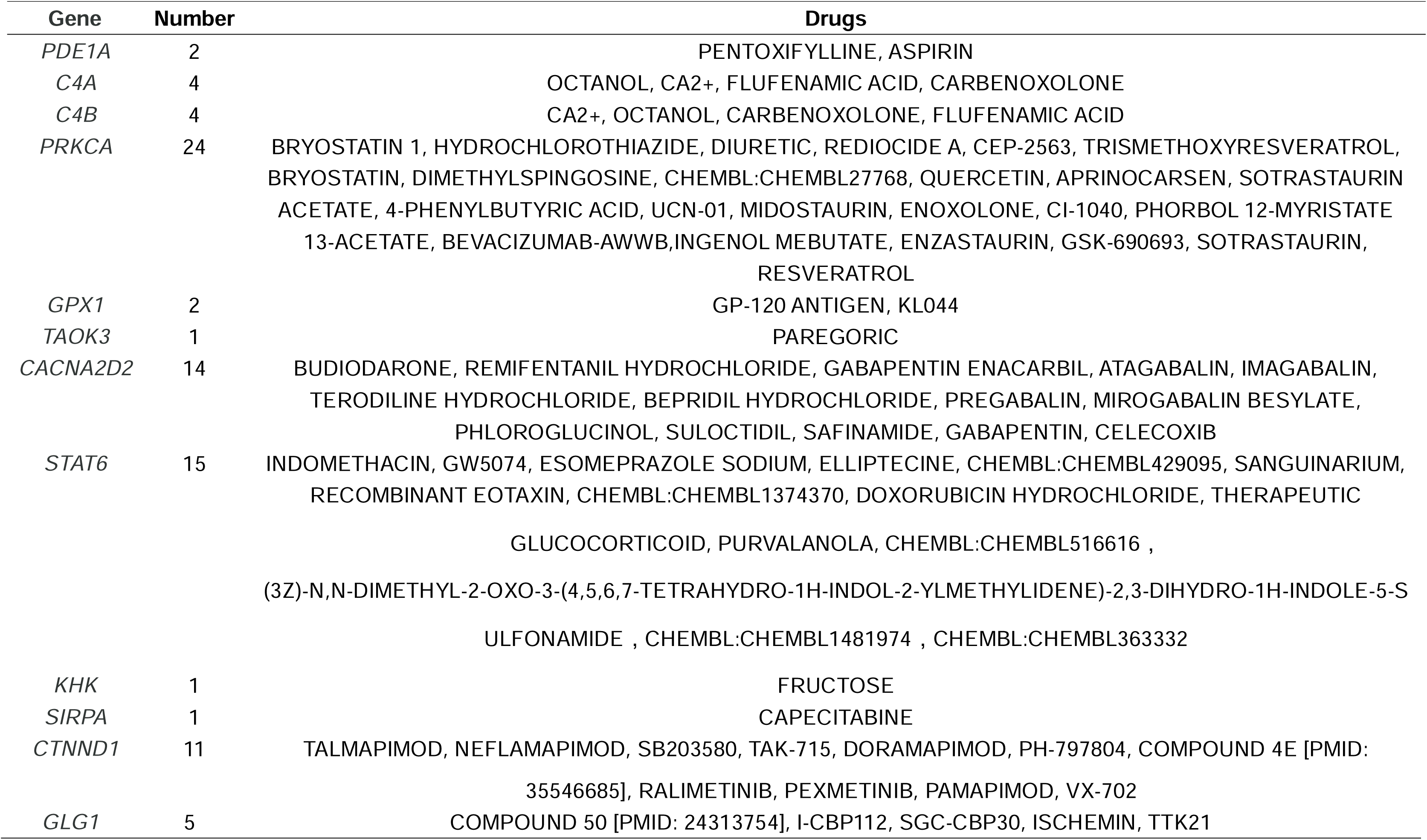
The drug-gene interactions of PWS genes from the DGIdb database.

### Gene expression and function analysis of the PWS genes in synapse

We queried the 33 PWS genes on SynGO website and found 5 genes, *RAB27B*, *PRKCA*, *CTNND1*, *CACNA2D2*, and *INPP4A* were annotated as function in synapse (**Figure 2**). *RAB27B, CACNA2D2, CTNND1,* and *PRKCA* were annotated as components of the presynapse. While *CTNND1* and INPP4A were annotated as components of the postsynapse. Noteblely, *CTNND1* was annotated in both the pre and postsynapse categories. The 33 PWS genes were found to be overrepresented in presynapse Cellular Component ontology (*P* = 0.04) and enriched in biological processes, including ‘regulation of postsynaptic membrane neurotransmitter receptor levels’ (*P* = 3.63×10^-03^) and ‘process in the synapse’ (*P*=0.05).

**Figure 2.**
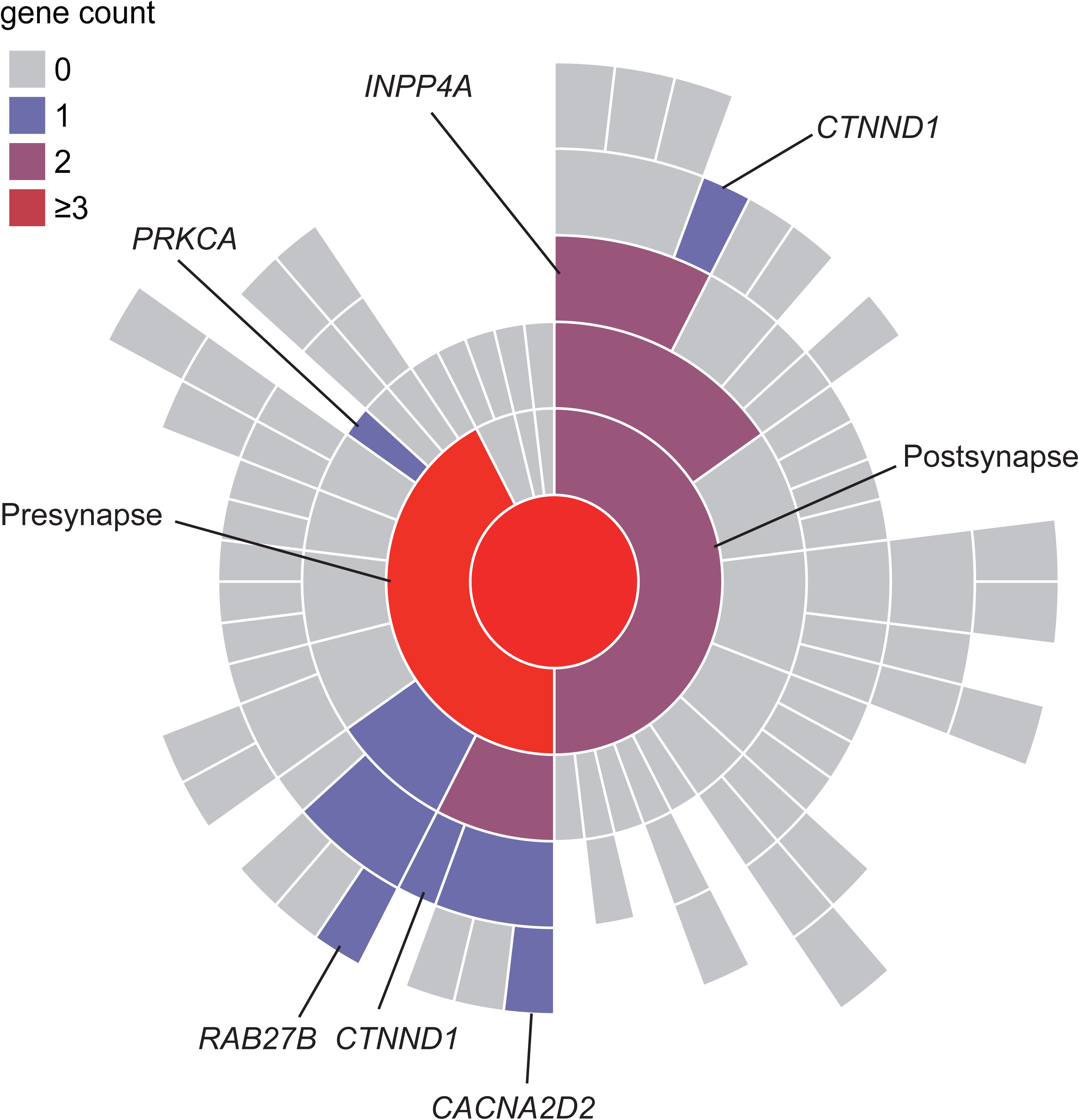
The Proteome-wide significant (PWS) genes that has annotation that related synapse by using SynGO web tool.

## Discussion

In this study, we performed PWAS analysis of PTSD GWAS summary statistics to identify potential protein abundance change that related to PTSD, ultimately identified 33 PWS proteins. Our findings expand our understanding of risk genes associated with PTSD. We compared our findings in this study with previous publications. Thomas et al. (2022), which conducted a PWAS analysis by integrating brain proteome and eight psychiatric traits [16]. They reported 16 PWS proteins that associated with PTSD. Of which 7 genes including *CTNND1*, *CCBL2*, *GPX1*, *INPP4A*, *KHK*, *PDLIM2*, and *RAB27B* were also identified in our study. Thomas et al. used a relatively small sample size PTSD GWAS summary statistics compared to this study, and they used a relaxed FDR adjustment for multiple comparison, while we employed a more stringent Bonferroni correction. Additionally, the newly released PGC-PTSD GWAS conducted a transcriptome-wide association study (TWAS) using GTEx brain tissue data, reporting 62 significant genes [12]. 6 genes overlapped with the PWS gene identified in our study, including *TMEM106B*, *GMPPB*, *KHK*, *GPX1*, *BTN3A3* and *C4A*. In summary, we identified 33 PWS genes in this study, providing several novel candidates that confer risk for PTSD and offering new molecular insights into the disease.

*CTNND1* (Catenin Delta 1) is our top finding in this study. It has been reported to function in cell-cell adhesion and play a role in neuronal systems pathways [17–19]. *CTNND1* has also been associated with depression, which is known to have a high comorbidity with PTSD [20–22]. Another gene C4A, was identified as a top association signal in schizophrenia and involved in synaptic function [23]. Our gene set enrichment analysis highlighted the 33 PWS genes are enriched in biological process including ‘regulation of postsynaptic membrane neurotransmitter receptor levels’ and ‘process in the synapse’. These evidences suggest that dysregulation of synapse function may contribute to the etiology of PTSD [24].

Our study has several limitations. Firstly, the proteome data we used was based entirely on human prefrontal cortex sample with a relatively small sample size. Further studies with larger sample size and data from multiple brain regions are warranted to identify additional risk genes for PTSD. Secondly, the PTSD GWAS cohort and brain proteome dataset utilized in this paper were predominantly from individuals of European ancestry. Future studies should include diverse ancestries to confirm our findings. Lastly, while our study aimed to identify risk genes associated with PTSD, the mechanisms through which these genes contribute to PTSD etiology require further investigation.

In summary, we performed PWAS analyses of PTSD by integrating the latest PTSD GWAS data with human brain proteome information. We identified a total of 33 PWS genes, which may serve as promising candidates for further studies on disease mechanisms and for the development of novel therapeutic drugs targeting these proteins.

## Data Availability

All data produced in the present study are available upon reasonable request to the authors

## Funding Statement

This study was funded by Hubei provincial Natural Science Foundation of China (2024AFB1022 to J.L) and the National Natural Science Foundation of China (U21A20364 to Z.L)

## Author contributions

JL and ZL convinced and supervised this study. JL performed the data analysis, visualization, and wrote the original draft. JL and ZL edited the manuscript.

## Conflict of Interest Disclosures

The author declared no Conflict of Interest

## Notes

### Competing Interest Statement

The authors have declared no competing interest.

### Author Declarations

The datasets used in this study are publicly available summary level data and were not generated as part of this research.

## References

1. Merians AN, Spiller T, Harpaz-Rotem I, Krystal JH, Pietrzak RH: Post-traumatic Stress Disorder. Med Clin North Am 2023, 107:85–99.

2. Koenen KC, Ratanatharathorn A, Ng L, McLaughlin KA, Bromet EJ, Stein DJ, Karam EG, Meron Ruscio A, Benjet C, Scott K, et al: Posttraumatic stress disorder in the World Mental Health Surveys. Psychol Med 2017, 47:2260–2274.

3. Davis LL, Schein J, Cloutier M, Gagnon-Sanschagrin P, Maitland J, Urganus A, Guerin A, Lefebvre P, Houle CR: The Economic Burden of Posttraumatic Stress Disorder in the United States From a Societal Perspective. J Clin Psychiatry 2022, 83.

4. Sartor CE, Grant JD, Lynskey MT, McCutcheon VV, Waldron M, Statham DJ, Bucholz KK, Madden PA, Heath AC, Martin NG, Nelson EC: Common heritable contributions to low-risk trauma, high-risk trauma, posttraumatic stress disorder, and major depression. Arch Gen Psychiatry 2012, 69:293–299.

5. Nievergelt CM, Maihofer AX, Klengel T, Atkinson EG, Chen CY, Choi KW, Coleman JRI, Dalvie S, Duncan LE, Gelernter J, et al: International meta-analysis of PTSD genome-wide association studies identifies sex- and ancestry-specific genetic risk loci. Nat Commun 2019, 10:4558.

6. Stein MB, Levey DF, Cheng Z, Wendt FR, Harrington K, Pathak GA, Cho K, Quaden R, Radhakrishnan K, Girgenti MJ, et al: Genome-wide association analyses of post-traumatic stress disorder and its symptom subdomains in the Million Veteran Program. Nat Genet 2021, 53:174–184.

7. Maihofer AX, Choi KW, Coleman JRI, Daskalakis NP, Denckla CA, Ketema E, Morey RA, Polimanti R, Ratanatharathorn A, Torres K, et al: Enhancing Discovery of Genetic Variants for Posttraumatic Stress Disorder Through Integration of Quantitative Phenotypes and Trauma Exposure Information. Biol Psychiatry 2022, 91:626–636.

8. Maurano MT, Humbert R, Rynes E, Thurman RE, Haugen E, Wang H, Reynolds AP, Sandstrom R, Qu H, Brody J, et al: Systematic localization of common disease-associated variation in regulatory DNA. Science 2012, 337:1190–1195.

9. Gallagher MD, Chen-Plotkin AS: The Post-GWAS Era: From Association to Function. Am J Hum Genet 2018, 102:717–730.

10. Gusev A, Ko A, Shi H, Bhatia G, Chung W, Penninx BW, Jansen R, de Geus EJ, Boomsma DI, Wright FA, et al: Integrative approaches for large-scale transcriptome-wide association studies. Nat Genet 2016, 48:245–252.

11. Zhu Z, Zhang F, Hu H, Bakshi A, Robinson MR, Powell JE, Montgomery GW, Goddard ME, Wray NR, Visscher PM, Yang J: Integration of summary data from GWAS and eQTL studies predicts complex trait gene targets. Nat Genet 2016, 48:481–487.

12. Nievergelt CM, Maihofer AX, Atkinson EG, Chen CY, Choi KW, Coleman JRI, Daskalakis NP, Duncan LE, Polimanti R, Aaronson C, et al: Genome-wide association analyses identify 95 risk loci and provide insights into the neurobiology of post-traumatic stress disorder. Nat Genet 2024, 56:792–808.

13. Wingo AP, Liu Y, Gerasimov ES, Gockley J, Logsdon BA, Duong DM, Dammer EB, Robins C, Beach TG, Reiman EM, et al: Integrating human brain proteomes with genome-wide association data implicates new proteins in Alzheimer’s disease pathogenesis. Nat Genet 2021, 53:143–146.

14. Cannon M, Stevenson J, Stahl K, Basu R, Coffman A, Kiwala S, McMichael JF, Kuzma K, Morrissey D, Cotto K, et al: DGIdb 5.0: rebuilding the drug-gene interaction database for precision medicine and drug discovery platforms. Nucleic Acids Res 2024, 52:D1227–d1235.

15. Koopmans F, van Nierop P, Andres-Alonso M, Byrnes A, Cijsouw T, Coba MP, Cornelisse LN, Farrell RJ, Goldschmidt HL, Howrigan DP, et al: SynGO: An Evidence-Based, Expert-Curated Knowledge Base for the Synapse. Neuron 2019, 103:217–234.e214.

16. Wingo TS, Liu Y, Gerasimov ES, Vattathil SM, Wynne ME, Liu J, Lori A, Faundez V, Bennett DA, Seyfried NT, et al: Shared mechanisms across the major psychiatric and neurodegenerative diseases. Nat Commun 2022, 13:4314.

17. Anastasiadis PZ, Moon SY, Thoreson MA, Mariner DJ, Crawford HC, Zheng Y, Reynolds AB: Inhibition of RhoA by p120 catenin. Nat Cell Biol 2000, 2:637–644.

18. Ishiyama N, Lee SH, Liu S, Li GY, Smith MJ, Reichardt LF, Ikura M: Dynamic and static interactions between p120 catenin and E-cadherin regulate the stability of cell-cell adhesion. Cell 2010, 141:117–128.

19. Park JI, Kim SW, Lyons JP, Ji H, Nguyen TT, Cho K, Barton MC, Deroo T, Vleminckx K, Moon RT, McCrea PD: Kaiso/p120-catenin and TCF/beta-catenin complexes coordinately regulate canonical Wnt gene targets. Dev Cell 2005, 8:843–854.

20. Liu J, Li X, Luo XJ: Proteome-wide Association Study Provides Insights Into the Genetic Component of Protein Abundance in Psychiatric Disorders. Biol Psychiatry 2021, 90:781–789.

21. Wingo TS, Liu Y, Gerasimov ES, Gockley J, Logsdon BA, Duong DM, Dammer EB, Lori A, Kim PJ, Ressler KJ, et al: Brain proteome-wide association study implicates novel proteins in depression pathogenesis. Nat Neurosci 2021, 24:810–817.

22. Flory JD, Yehuda R: Comorbidity between post-traumatic stress disorder and major depressive disorder: alternative explanations and treatment considerations. Dialogues Clin Neurosci 2015, 17:141–150.

23. Sekar A, Bialas AR, de Rivera H, Davis A, Hammond TR, Kamitaki N, Tooley K, Presumey J, Baum M, Van Doren V, et al: Schizophrenia risk from complex variation of complement component 4. Nature 2016, 530:177–183.

24. Krystal JH, Abdallah CG, Averill LA, Kelmendi B, Harpaz-Rotem I, Sanacora G, Southwick SM, Duman RS: Synaptic Loss and the Pathophysiology of PTSD: Implications for Ketamine as a Prototype Novel Therapeutic. Curr Psychiatry Rep 2017, 19:74.

